# Genetic Evidence for Opposing Associations of Psoriasis and Type 2 Diabetes with Inflammatory Bowel Disease: A Mendelian Randomization Study

**DOI:** 10.64898/2026.02.25.26346967

**Authors:** Mathilde Riger Orkild, Karoline Lindgaard Dybdahl, Palle Duun Rohde

## Abstract

Inflammatory bowel disease (IBD) frequently co-occurs with immune-mediated and metabolic disorders, but whether these associations reflect shared genetics or causal effects remains unclear. We performed two-sample Mendelian randomization (MR) using large-scale genome-wide association study (GWAS) summary statistics to investigate potential causal effects of immune-mediated diseases and lifestyle traits on IBD, Crohn’s disease (CD), and ulcerative colitis (UC). SNP-based heritability and genetic correlations were estimated to contextualize findings. Following false discovery rate correction, genetically predicted psoriasis was positively associated with IBD (OR 1.15), CD (OR 1.23), and UC (OR 1.10), with the strongest effect observed for CD. Genetically predicted type 2 diabetes mellitus (T2DM) showed a modest inverse association with UC (OR 0.88). No lifestyle-related traits remained significant after correction. Sensitivity analyses indicated heterogeneity across instruments and evidence of directional pleiotropy in selected models, whereas no pleiotropy was detected for the T2DM–UC association. These findings support a role of psoriasis-related immune pathways in IBD susceptibility and suggest a potential inverse association between genetic liability to T2DM and UC.

## Introduction

Inflammatory bowel diseases (IBD) are chronic, immune-mediated inflammatory disorders of the gastrointestinal (GI) tract that represent a rapidly increasing global health burden (Din et al., 2020; Guan, 2019; Wang et al., 2016). Affecting more than 0.3% of the global population (Desai & Dhoble, 2024), IBD prevalence has increased by over 45% between 1990 and 2019 (Wang et al., 2023). The disease is associated with substantial morbidity and a threefold increase in annual healthcare costs in the United States compared with unaffected individuals (Park et al., 2019). Despite advances in medical therapy – including aminosalicylates, corticosteroids, biologics, and surgery – IBD remains incurable and may lead to severe complications such as strictures, fistulas (Agrawal et al., 2021), and colorectal cancer (McDowell et al., 2023), thereby intensifying pressure on healthcare systems (Wang et al., 2023).

IBD comprises two principal subtypes: ulcerative colitis (UC) and Crohn’s disease (CD) (Cleveland Clinic, 2024; Din et al., 2020; Guan, 2019; Zhang & Li, 2014). While UC is characterized by continuous mucosal inflammation restricted to the colon, CD involves discontinuous, transmural inflammation that can affect any segment of the GI tract (Guan, 2019; Zhang & Li, 2014). Although clinically overlapping, these subtypes differ in anatomical distribution, immune activation patterns, and complication profiles.

The aetiology of IBD is multifactorial and involves impaired epithelial barrier integrity, microbial dysbiosis, aberrant immune activation, and genetic susceptibility (Guan, 2019; McDowell et al., 2023; Wang et al., 2023; Zhang & Li, 2014). Large-scale genome-wide association studies (GWAS) have identified numerous susceptibility loci implicating pathways central to immune regulation and intestinal homeostasis (De Lange et al., 2017; Guan, 2019; Liu et al., 2023; Mehta et al., 2017; Zhu et al., 2025). Many of these loci converge on immune signalling networks, including pathways regulating T-cell differentiation and cytokine signalling, suggesting biological overlap with other immune-mediated diseases.

Indeed, IBD frequently co-occurs with autoimmune and metabolic disorders such as psoriasis (PSO), rheumatoid arthritis (RA), myasthenia gravis (MG), type 1 diabetes mellitus (T1DM), and type 2 diabetes mellitus (T2DM) (Kumar et al., 2021; Mosli et al., 2022). Several of these conditions share pathogenic mechanisms characterized by dysregulated adaptive immunity and loss of self-tolerance (Pisetsky, 2023; Shoenfeld & Isenberg, 1989). In particular, psoriasis and IBD exhibit partially overlapping genetic risk loci and immune signatures, including involvement of the IL-23/Th17 axis, raising the possibility of shared upstream inflammatory drivers rather than simple comorbidity (Gaffen et al., 2014; Jostins et al., 2012; Nair et al., 2009). However, whether such overlap reflects true directional causal effects or merely shared polygenic susceptibility remains unresolved.

At the same time, GWAS demonstrate that IBD shares susceptibility loci with several immune-mediated diseases, including psoriasis, within key inflammatory pathways such as IL23R, TYK2 and STAT3. Significant genome-wide genetic correlation and non-trivial SNP-based heritability estimates further support partially overlapping polygenic architecture (Gaffen et al., 2014; Jostins et al., 2012; Nair et al., 2009). Yet shared heritability is agnostic to directionality and may reflect horizontal pleiotropy or common upstream inflammatory programs rather than direct causal relationship (van Rheenen et al., 2019). Together, these epidemiological and genetic observations underscore the difficulty of distinguishing true causal drivers from correlates of shared biology.

Lifestyle-related traits further complicate this landscape. Western dietary patterns characterized by high intake of processed foods, salt, and refined carbohydrates, together with reduced fruit and vegetable consumption (Cordain et al., 2005; Rizzello et al., 2019), as well as sedentary behaviour and reduced physical activity(Adolph & Tilg, 2024), have been associated with IBD risk (Lopes et al., 2023; Piovani et al., 2019; Rizzello et al., 2019). Smoking and coffee intake have similarly been implicated (Mahid et al., 2006; Parkes et al., 2014). However, observational associations between lifestyle exposures and IBD are vulnerable to residual confounding, reverse causation, and behavioural modification following subclinical disease onset, complicating causal interpretation (Smith & Hemani, 2014).

MR provides a framework for strengthening causal inference by leveraging germline genetic variants associated with exposures as instrumental variables (Boehm & Zhou, 2022; Davies et al., 2018). Because genetic variants are randomly allocated at conception and fixed throughout life, MR reduces bias from confounding and reverse causation. Large publicly available GWAS datasets enable two-sample MR analyses across diverse traits (Tam et al., 2019), offering a scalable complement to randomized controlled trials, which may be infeasible, unethical, or impractical for many exposures (Hariton & Locascio, 2018; Teumer, 2018). Valid inference requires that instruments satisfy assumptions of relevance, independence, and exclusion restriction (Bowden et al., 2015).

In this study, we performed a large-scale two-sample MR analysis to systematically evaluate whether five immune-mediated or metabolic diseases and twelve genetically proxied lifestyle-related traits exert causal effects on IBD and its subtypes, UC and CD. By integrating SNP-based heritability estimation, pairwise genetic correlation analyses, and rigorously controlled two-sample MR, we sought to disentangle shared polygenic liability from directional causal effects. Clarifying whether comorbid immune and metabolic disorders represent true upstream drivers of IBD has important implications for mechanistic understanding, risk stratification, and identification of potentially modifiable pathways in a complex inflammatory disease.

## Method

### Study Design and Core Assumptions of Mendelian Randomization

The overall study workflow is illustrated in Figure 1.

**Figure 1.**
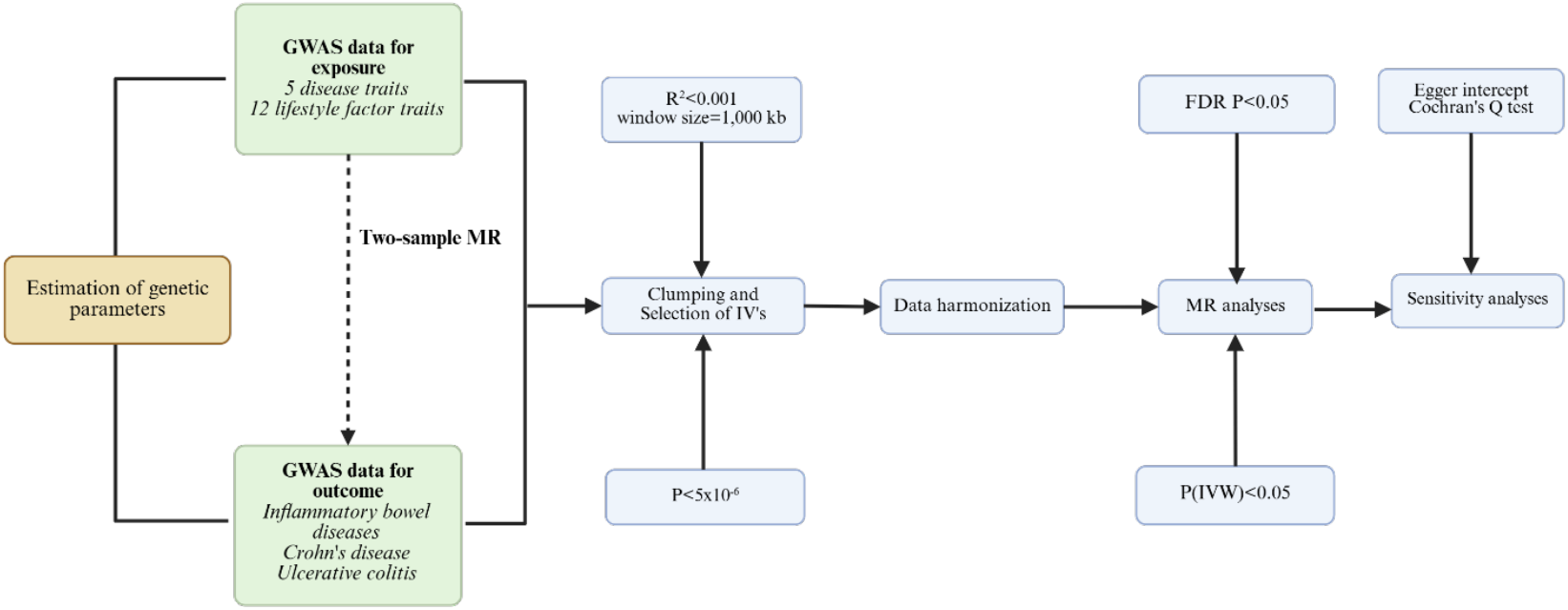
Overview of the two-sample Mendelian randomization workflow. The analytical pipeline illustrates the selection of genetic instruments from GWAS summary statistics for five disease traits and twelve lifestyle-related traits. Prior to Mendelian randomization (MR), SNP-based heritability (h^2^) and genetic correlation (r_g_) between exposures and outcomes were estimated. Genetic instruments were selected using genome-wide significance thresholds (P < 5×10^−6^) and linkage disequilibrium (LD) clumping (r^2^ < 0.001, window size = 1,000 kb). Harmonized exposure and outcome datasets were subsequently used to perform inverse-variance weighted (IVW) MR analyses across inflammatory bowel disease (IBD), Crohn’s disease (CD), and ulcerative colitis (UC). Multiple testing was controlled using false discovery rate (FDR) correction (FDR-adjusted P < 0.05). Sensitivity analyses included assessment of horizontal pleiotropy and heterogeneity using the MR-Egger intercept and Cochran’s Q statistic, respectively. Abbreviations: MR, Mendelian randomization; GWAS, genome-wide association study; IV, instrumental variable; LD, linkage disequilibrium; LDSC, linkage disequilibrium score regression; IVW, inverse-variance weighted; FDR, false discovery rate.

A two-sample MR design was employed to estimate causal associations using publicly available GWAS summary statistics (Davies et al., 2018). The analytical framework and core assumptions underlying the MR analyses are depicted in Figure 2.

**Figure 2.**
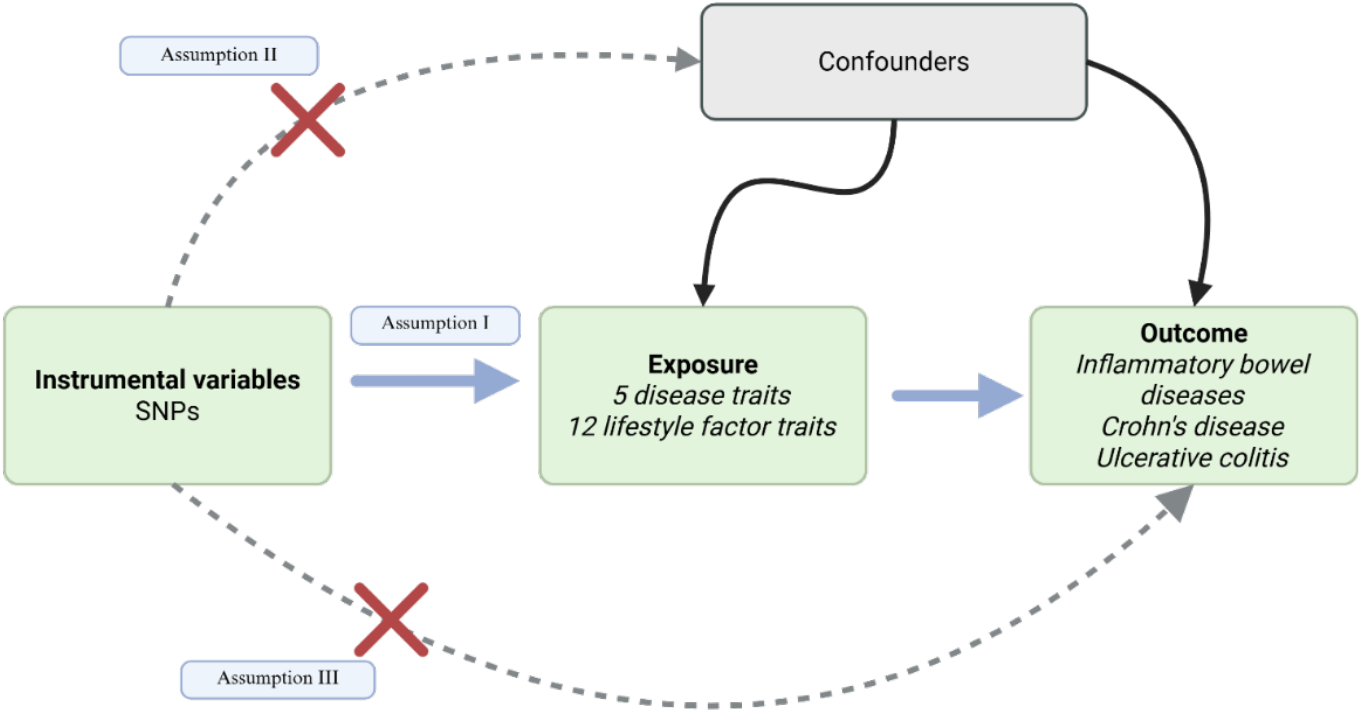
Conceptual illustration of the three core assumptions underlying Mendelian randomization. The diagram depicts the instrumental variable framework applied in this study. Assumption I (relevance): genetic instruments (SNPs) are robustly associated with the exposure. Assumption II (independence): instruments are independent of confounders of the exposure–outcome relationship. Assumption III (exclusion restriction): instruments influence the outcome only through the exposure and not via alternative pathways (i.e., absence of horizontal pleiotropy). Arrows represent hypothesized causal pathways, while dashed lines with crosses indicate prohibited associations under valid instrumental variable assumptions. Abbreviations: MR, Mendelian randomization; SNPs, single-nucleotide polymorphisms.

MR relies on three fundamental assumptions:

1. Relevance (Assumption I): The instrumental variables (IVs), here single-nucleotide polymorphisms (SNPs), must be robustly associated with the exposure of interest.
2. Independence (Assumption II): The IVs must be independent of confounders of the exposure–outcome relationship.
3. Exclusion Restriction (Assumption III): The IVs must influence the outcome exclusively through the exposure and not via alternative pathways.Violation of these assumptions may bias causal estimates.

### Exposure and Outcome Definition

Two sample MR analyses were conducted to estimate the causal association between selected immune-mediated diseases or lifestyle-related traits and the IBD spectrum (IBD overall, CD, and UC). Six diseases were included to represent a spectrum of autoimmune and metabolically relevant conditions: myasthenia gravis, psoriasis, rheumatoid arthritis, T1DM, and T2DM. In addition, twelve lifestyle-related traits were included, comprising dietary patterns, anthropometric traits, smoking-related variables, and behavioural measures (e.g., processed meat consumption, pack-years, sugar intake). These traits were selected to evaluate whether genetic liability to lifestyle tendencies is causally associated with IBD risk.

Across five diseases and twelve lifestyle traits evaluated against three IBD outcomes, a total of 51 exposure– outcome pairs were analysed.

### Data sources

GWAS summary statistics were obtained from the GWAS Catalogue (Cerezo et al., 2025). Detailed information for all included datasets is provided in Supplementary Table S1. The majority of GWAS datasets were derived from individuals of European ancestry. Restricting analyses to predominantly European ancestry was intended to minimise bias arising from differences in allele frequencies and linkage disequilibrium (LD) patterns across populations (Davies et al., 2018).

GWAS summary statistics for multifactorial complex diseases were used as exposure variables in the MR analyses. The following disease traits were included: myasthenia gravis (N = 437,736), psoriasis (N = 494,544), rheumatoid arthritis (N = 97,173), T1DM (N = 173,981), and T2DM (N = 298,957). To evaluate potential causal effects of genetically proxied lifestyle-related tendencies, GWAS summary statistics were additionally obtained for twelve behavioural and dietary traits: added salt consumption (N = 448,890), coffee consumption (N = 448,204), fresh fruit intake (N = 447,401), leisure sedentary behaviour (N = 408,815), pack-years of smoking (N = 377,234), processed meat consumption (N = 448,303), smoking behaviour (N = 518,633), sugar consumption (N = 447,391), carbohydrate intake (N = 282,271), protein intake (N = 282,271), and fat intake (N = 282,271). All sample sizes correspond to those reported in the original GWAS publications.

### Estimation of SNP-Based Heritability

SNP-based heritability 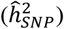 was estimated for all included traits using the same GWAS summary statistics as those applied in the MR analyses. 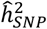 quantifies the proportion of phenotypic variance attributable to additive genetic variation tagged by common SNPs. Estimates range from 0 to 1, where 0 indicates no contribution from common genetic variants and 1 indicates that all phenotypic variance is attributable to such variants within the studied population (Speed et al., 2017; Speed & Balding, 2019). Heritability estimates were calculated using SumHer implemented in the BLD-LDAK software (Speed & Balding, 2019. Estimates are reported with corresponding standard errors (SE).

### Estimation of Genetic Correlation

Pairwise genetic correlations 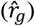 between IBD outcomes (IBD, CD, UC) and each exposure trait were estimated using the same GWAS summary statistics applied in the MR analyses. 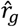 were calculated using SumHer (Speed & Balding, 2019) with LDAK-Thin (version 6.1) as the tagging file. The 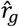 reflects the correlation of SNP effect sizes between two traits and ranges from −1 to 1. Positive values indicate concordant allelic effects, whereas negative values indicate discordant effects (Pal & Chakravarty, 2020). Estimates are reported with 95% confidence intervals (CIs). If the CI included zero, the 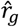 indistinguishable from zero. was considered statistically

### Instrument Selection and Quality Control

A series of quality control procedures was applied to exposure GWAS summary statistics prior to MR analysis. First, SNPs associated with each exposure at *P*<1×10^−5^ were selected as candidate instrumental variables. Second, linkage disequilibrium (LD) clumping was performed to ensure independence between instruments. Strong LD (measured as Pearson’s r^2^) may induce biased causal estimates (Boehm & Zhou, 2022; Davies et al., 2018). Clumping was conducted using a European reference panel with the following parameters: *r*^2^ threshold <0.001, window size 10,000 kb, and threshold of *P* < 5 × 10^−6^. SNPs meeting these criteria were retained for MR analysis.

### Harmonisation

Harmonisation of exposure and outcome summary statistics was performed to ensure consistent alignment of effect alleles across datasets. Ambiguous or palindromic SNPs with incompatible allele frequencies were excluded. Where necessary, allele orientation was corrected to ensure that effect estimates corresponded to the same effect allele prior to MR estimation.

### Mendelian Randomization Analysis

Causal effects were estimated using the inverse-variance weighted (IVW) method, producing 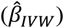 estimates for each exposure–outcome pair. The IVW method assumes that all included SNPs are valid instruments and provides the highest statistical power under this assumption; however, it may be biased in the presence of invalid instruments or directional pleiotropy (Burgess et al., 2015). To evaluate robustness of causal estimates, complementary sensitivity analyses were performed using the weighted median and MR-Egger methods. The MR-Egger intercept was used to assess directional pleiotropy. Heterogeneity across instruments was assessed using Cochran’s Q statistic. To account for multiple testing, false discovery rate (FDR) correction (Dunn, 1961; Glickman et al., 2014) was applied separately within each outcome (IBD, CD, UC) and within exposure categories (disease traits and lifestyle traits). FDR-adjusted *P*-values < 0.05 were considered statistically significant.

Causal estimates are reported as odds ratios (ORs) with corresponding 95% CIs. All MR analyses were conducted using the “TwoSampleMR” package in R (R Core Team, 2021).

## Results

### SNP-based Heritability and Genetic Correlations of Diseases and Lifestyle Factors

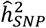 was estimated to quantify the proportion of phenotypic variance attributable to common genetic variation for each trait. Estimates and standard errors are presented in Figure 3 and detailed in Supplementary Table S2. All estimates were derived from GWAS summary statistics used in subsequent MR analyses. Across all traits 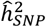 ranged from 0.018±0.002 to 0.272±0.011. Most traits demonstrated modest heritability estimates below 0.30. For the outcome traits (IBD, CD, and UC; Figure 3A), 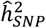 ranged from 0.099 (±0.003) to 0.127 (±0.003). Among disease exposure traits (Figure 3B), estimates ranged from 0.018 (±0.002) to 0.272 (±0.011). Lifestyle factor traits (Figure 3C) showed 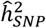 estimates between 0.030 (±0.003) and 0.180 (±0.003).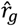 were estimated between each exposure trait and IBD, CD, and UC using LD score regression (Supplementary Table S3). Correlations between disease traits and outcomes (Figure 3D) ranged from –0.106 to 0.262. For lifestyle factor traits (Figure 3E), 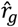 values ranged from –0.062 to 0.127.

**Figure 3.**
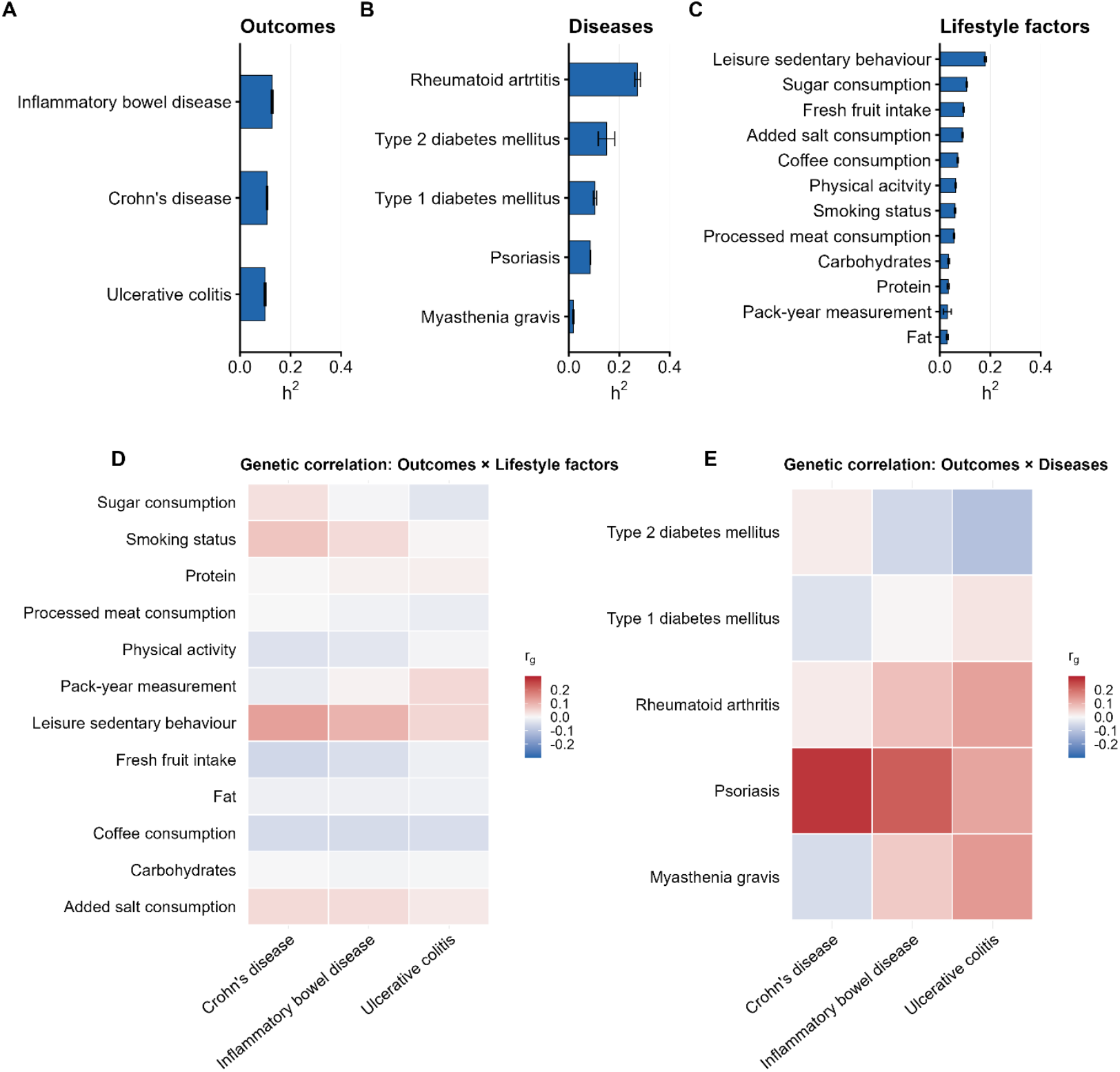
SNP-based heritability and genetic correlations of disease and lifestyle traits with inflammatory bowel disease outcomes. (A–C) Bar plots showing SNP-based heritability (*h*^2^) estimates with standard errors for (A) outcome traits (inflammatory bowel disease [IBD], Crohn’s disease [CD], and ulcerative colitis [UC]), (B) disease exposure traits, and (C) lifestyle factor traits. Heritability estimates were calculated from GWAS summary statistics using the SumHer approach. Error bars represent standard errors of the estimated *h*^2^. (D–E) Heatmaps displaying genetic correlations (r_g_) between exposure traits and outcomes. (D) Genetic correlations between lifestyle factor traits and IBD, CD, and UC. (E) Genetic correlations between disease traits and IBD, CD, and UC. Colour intensity reflects the magnitude and direction of the genetic correlation, with red indicating positive correlations and blue indicating negative correlations. Abbreviations: *h*^2^, SNP-based heritability; r_g_, genetic correlation; GWAS, genome-wide association study; IBD, inflammatory bowel disease; CD, Crohn’s disease; UC, ulcerative colitis.

### Identification of Genetic Causal Association Between Diseases or Lifestyle Factors and Inflammatory Bowel Diseases

Two-sample MR analyses were performed using the IVW method based on GWAS summary statistics. After FDR correction, four associations within the disease exposure traits remained statistically significant (Figure 4, Supplementary Table S4, S6). Genetically predicted psoriasis was positively associated with all three outcomes: CD (OR 1.22, 95% CI 1.12–1.34, *P*_FDR_ < 0.001), IBD (OR 1.15, 95% CI 1.07–1.23, *P*_FDR_ < 0.001), and UC (OR 1.10, 95% CI 1.03–1.18, *P*_FDR_ = 0.033). Genetically predicted T2DM was inversely associated with UC (OR 0.88, 95% CI 0.80–0.97, *P*_FDR_ = 0.041). MR-Egger intercept tests indicated evidence of directional horizontal pleiotropy for several exposure–outcome pairs (Supplementary Table S10). After FDR correction, significant intercepts were observed for myasthenia gravis in relation to CD and IBD, and for psoriasis and rheumatoid arthritis in relation to IBD. A significant intercept was also observed for psoriasis in relation to UC. No evidence of directional pleiotropy was detected for the association between genetically predicted T2DM and UC (*P*_intercept_ > 0.05, Supplementary Table S10). Cochran’s Q tests indicated substantial heterogeneity across instrumental variables for most exposure–outcome pairs (all *P*_Q_ < 0.001 after FDR correction; Supplementary Table S8), suggesting variability in SNP-specific causal estimates. Together with the MR-Egger intercept results, these findings indicate potential pleiotropic effects in several models and warrant cautious interpretation of the corresponding associations. No lifestyle factor traits remained statistically significant after FDR correction (Figure 4-5; Supplementary Table S5, S7, S11).

**Figure 4.**
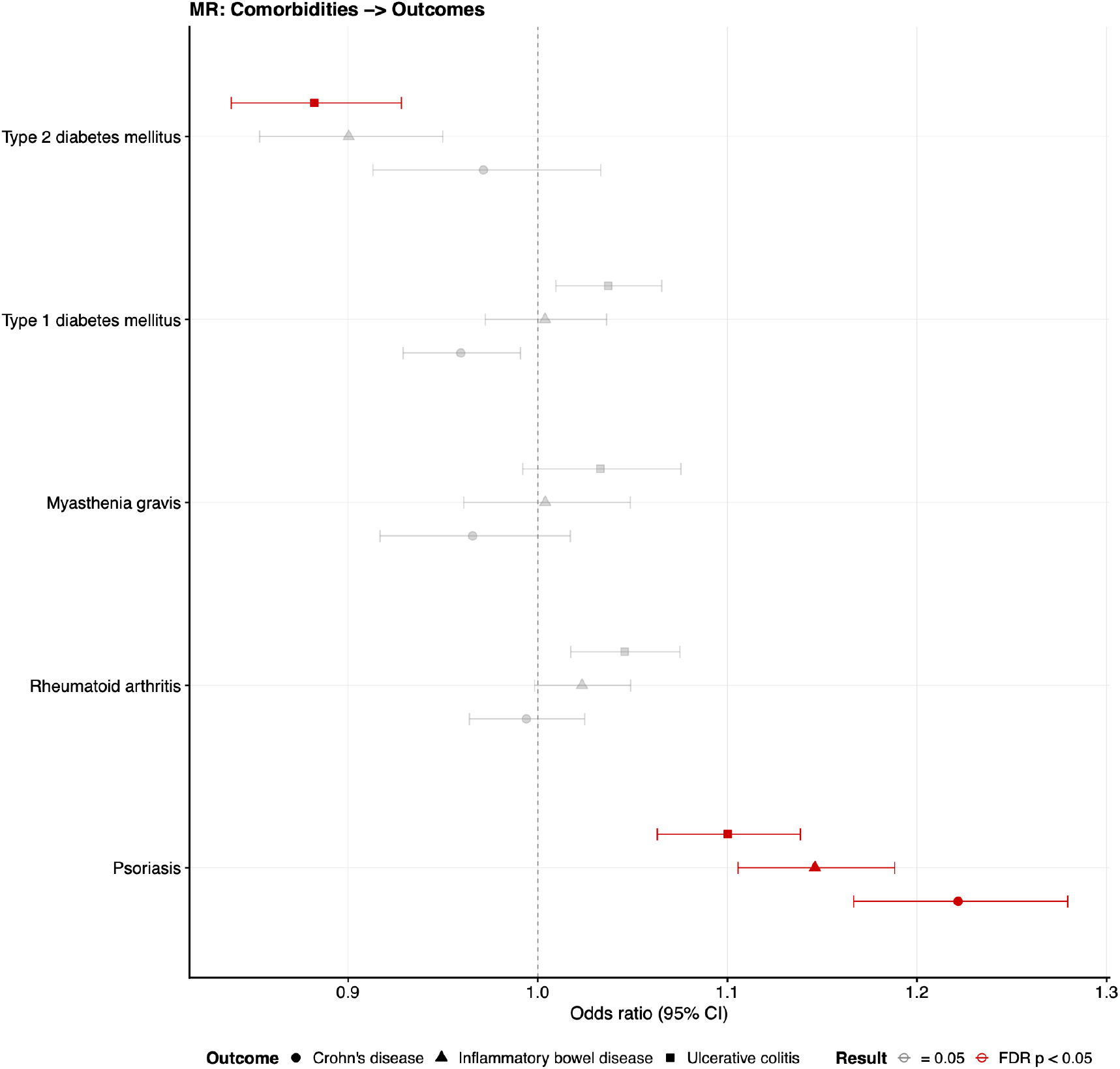
Mendelian randomization estimates for disease traits in relation to inflammatory bowel disease outcomes. Forest plots showing inverse-variance weighted (IVW) causal effect estimates from two-sample Mendelian randomization analyses of. Disease traits were evaluated for their associations with Crohn’s disease (circles), inflammatory bowel disease (triangles), and ulcerative colitis (squares). Effect estimates are presented as odds ratios (ORs) with 95% confidence intervals (CIs). The vertical dashed line indicates the null value (OR = 1). False discovery rate (FDR) correction was applied to account for multiple testing. Red markers indicate associations remaining statistically significant after FDR correction (*P*_FDR_ < 0.05). Black markers indicate nominal significance (95% CI excluding 1.0 but *P*_FDR_ ≥ 0.05), and grey markers indicate non-significant associations. Abbreviations: MR, Mendelian randomization; IVW, inverse-variance weighted; OR, odds ratio; CI, confidence interval; FDR, false discovery rate.

**Figure 5.**
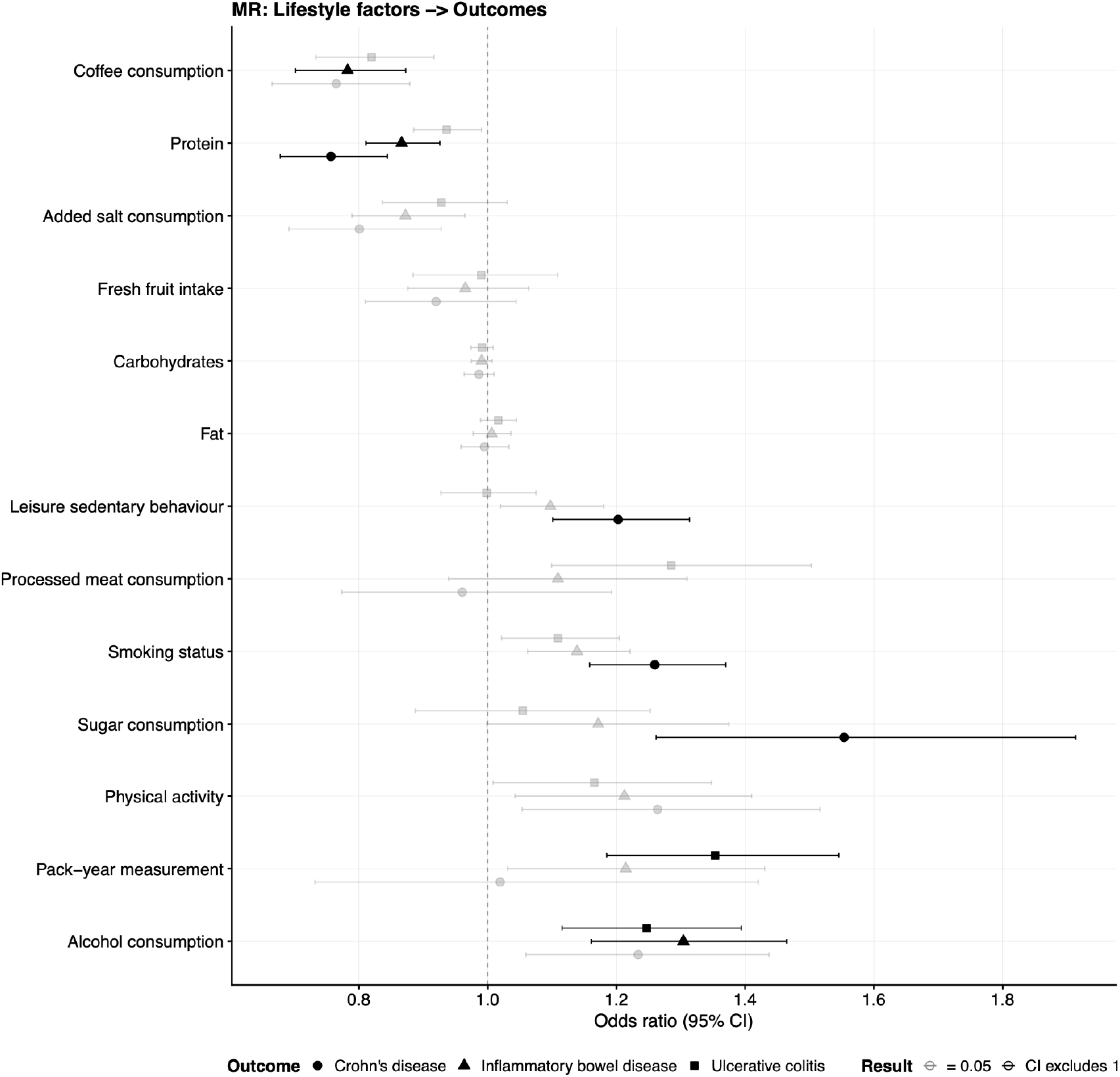
Mendelian randomization estimates for lifestyle factor traits in relation to inflammatory bowel disease outcomes. Forest plots showing inverse-variance weighted (IVW) causal effect estimates from two-sample Mendelian randomization analyses. Lifestyle factor traits were evaluated for their associations with Crohn’s disease (circles), inflammatory bowel disease (triangles), and ulcerative colitis (squares). Effect estimates are presented as odds ratios (ORs) with 95% confidence intervals (CIs). The vertical dashed line indicates the null value (OR = 1). False discovery rate (FDR) correction was applied to account for multiple testing. Red markers indicate associations remaining statistically significant after FDR correction (*P*_FDR_ < 0.05). Black markers indicate nominal significance (95% CI excluding 1.0 but *P*_FDR_ ≥ 0.05), and grey markers indicate non-significant associations. Abbreviations: MR, Mendelian randomization; IVW, inverse-variance weighted; OR, odds ratio; CI, confidence interval; FDR, false discovery rate.

In addition to the FDR-significant findings, several exposure–outcome associations demonstrated nominal statistical significance (i.e., 95% CI excluding 1.0) but did not remain significant after FDR correction (Figure 4-5; Supplementary Table S4–S5). These associations should be interpreted cautiously, as they did not withstand adjustment for multiple testing. No disease traits demonstrated nominal associations except for the four FDR-significant findings already observed. Among the lifestyle factor traits, several estimates showed CIs excluding the null value: Protein consumption on CD (OR 0.76, 95% CI 0.61-0.94) and IBD (OR 0.87 95% CI 0.76-0.99), leisure sedentary behaviour on CD (OR 1.20, 95% CI 1.01-1.43), sugar consumption on CD (OR 1.55, 95% CI 1.03-2.34), smoking status on CD (OR 1.26, 95% CI 1.07-1.49), Coffee consumption on IBD (OR 0.78, 95% CI 0.63-0.97), and pack-year-measurement on UC (OR 1.35, 95% CI 1.04-1.76). All of these results indicate potential associations at the nominal level, but none remained statistically significant following multiple testing correction.

## Discussion

Using large-scale GWAS summary statistics, we applied two-sample MR to investigate potential causal effects of immune-mediated diseases and lifestyle-related traits on IBD, including CD and UC. In addition, 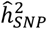 and genome-wide 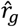 were estimated to contextualize the MR findings.

Across traits, 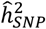 estimates were generally modest, consistent with the multifactorial and polygenic nature of both immune-mediated diseases and lifestyle behaviours. Genetic correlations between exposures and IBD outcomes were overall small to moderate, with most CIs overlapping zero. These findings indicate limited genome-wide shared polygenic architecture for most traits and reinforce that genetic correlation does not imply causality.

Importantly, several traits with minimal or non-significant genetic correlation still showed suggestive or significant MR estimates, highlighting that causal inference from MR is not dependent on strong genome-wide correlation.

The most robust finding of this study was a positive causal association between genetically predicted psoriasis and all three outcomes (IBD, CD, and UC), with the strongest effect observed for CD. MR-Egger intercept analyses indicated evidence of directional pleiotropy in the models for overall IBD and UC, whereas no significant intercept was observed for CD after multiple testing correction. These findings suggest that shared immune-related genetic architecture may partly contribute to the associations with IBD and UC. Nevertheless, the consistent direction of effect across outcomes and concordance with previous MR studies support a potential causal role of psoriasis-related pathways in IBD susceptibility. Sun et al. (2022) reported a significant causal effect of psoriasis on CD but not UC (Sun et al., 2022), while Cai et al. (2023) observed an effect on overall IBD (Cai et al., 2023). Observational data have also documented bidirectional comorbidity between psoriasis and IBD (Bernstein et al., 2005; Fu et al., 2018). Biologically, psoriasis and IBD share susceptibility loci (Ellinghaus et al., 2012; Skroza et al., 2013) and immune pathways, particularly involving the IL-23/Th17 axis (Bianchi & Rogge, 2019; Maddur et al., 2012; McGeachy et al., 2009). Elevated IL-17 levels have been demonstrated in both psoriasis (Chan et al., 2006) and IBD (Fujino et al., 2003), supporting overlapping inflammatory mechanisms. The moderate genetic correlation observed between psoriasis and CD further supports shared biology, although MR provides evidence that this overlap may translate into a directional causal influence rather than mere polygenic co-occurrence. Collectively, these results strengthen evidence that psoriasis-related immune dysregulation may contribute to IBD susceptibility, particularly CD.

We observed a modest inverse association between genetically predicted T2DM and UC. No significant associations were observed for CD or overall IBD after multiple testing correction. This finding aligns with Xu et al. and Xiao et al., who also reported inverse associations between genetically predicted T2DM and UC (Xiao et al., 2024; Xu et al., 2024), whereas Tang et al. found no association (Tang et al., 2024). The underlying biological explanation remains unclear. Genetic correlation between T2DM and UC was close to zero, suggesting limited shared polygenic architecture. The observed inverse MR estimate therefore likely reflects pathway-specific mechanisms rather than broad genetic overlap. It is important to emphasize that MR estimates reflect the effect of lifelong genetic liability to T2DM, not the clinical or metabolic consequences of overt diabetes. Observational studies, including a Danish cohort study (Jess et al., 2020), have reported increased T2DM incidence among patients with IBD, highlighting that reverse causation, inflammation, medication effects, and lifestyle factors may influence epidemiological associations differently from genetically proxied liability. Thus, the present findings should be interpreted as evidence that genetic predisposition to T2DM-related pathways may modestly reduce UC susceptibility, rather than suggesting a clinically protective effect of diabetes itself.

For the remaining disease traits, no consistent causal associations were observed after multiple testing correction. Some nominal associations were noted but did not withstand correction. Previous MR studies have reported heterogeneous findings across the included non-significant disease traits and IBD, including T1DM (Tong et al., 2024; Xie et al., 2024; J. Y. Zhu et al., 2024). Differences across studies may reflect variation in GWAS datasets, ancestry composition, instrument selection thresholds, or statistical power. Our results suggest that, beyond psoriasis, there is limited robust evidence for strong causal effects of other investigated diseases on IBD susceptibility in this dataset.

No lifestyle-related traits remained statistically significant after multiple testing correction. However, several exposures showed nominal associations (e.g., pack-years, smoking status, and coffee consumption) with CIs excluding unity but not surviving FDR correction. Previous MR studies have reported modest effects of smoking on IBD risk (Carreras-Torres et al., 2020; Jones et al., 2020), consistent with established epidemiological evidence (Mahid et al., 2006). Our findings neither strongly confirm nor definitively refute these associations but suggest that genetically proxied behavioural traits explain only a limited proportion of IBD susceptibility. For dietary traits characteristic of a Western diet (Cordain et al., 2005; Rizzello et al., 2019), no causal effects were observed, in agreement with previous MR analyses (Chen et al., 2022; Wang et al., 2024). These findings suggest that observed epidemiological associations between diet and IBD may involve non-genetic environmental mechanisms or measurement heterogeneity rather than shared genetic liability.

A major strength of this study is the use of large GWAS datasets and systematic evaluation across immune-mediated and lifestyle exposures. Sensitivity analyses identified evidence of directional pleiotropy in several exposure–outcome pairs, particularly for psoriasis and certain immune-mediated traits in relation to overall IBD. These findings highlight the importance of cautious interpretation, as some associations may reflect shared biological pathways rather than purely vertical causal effects. Notably, no evidence of directional pleiotropy was observed for the inverse association between genetically predicted T2DM and UC, supporting the robustness of this finding. Cochran’s Q tests indicated substantial heterogeneity across instrumental variables in most analyses. Such heterogeneity is common in polygenic traits and may reflect biological complexity, variability in SNP-specific pathways, or horizontal pleiotropy. While heterogeneity does not invalidate MR assumptions per se, it underscores that causal estimates represent average effects across genetically heterogeneous instruments. The presence of heterogeneity, together with evidence of directional pleiotropy in selected models, warrants cautious interpretation of specific exposure–outcome associations.

Limitations include partial ancestry heterogeneity in some GWAS datasets, which may introduce residual population stratification. Differences in instrument selection thresholds, LD reference panels, and case definitions across studies may also contribute to discrepancies with prior MR findings. Additionally, MR estimates reflect lifelong genetic liability and cannot capture time-varying exposures or disease severity.

This MR study provides evidence that genetically predicted psoriasis increases susceptibility to IBD, particularly Crohn’s disease, while genetically predicted T2DM is modestly inversely associated with ulcerative colitis. No robust causal effects were observed for the remaining immune-mediated diseases or lifestyle traits after correction for multiple testing. These findings refine our understanding of shared immune and metabolic pathways in IBD pathogenesis and underscore the importance of distinguishing genetic correlation from causal inference in complex immune-mediated diseases.

## Data availability

The genome-wide association study (GWAS) summary statistics used in this study are publicly available from the original consortia and repositories cited in the References. Specific accession numbers, URLs, and study details are provided in Supplementary Table S1. All data analyzed in this study are derived from publicly available summary-level datasets. No individual-level data were accessed.

## Ethics Approval and Consent to Participate

This study used publicly available, de-identified GWAS summary statistics. All original studies received ethical approval from their respective institutional review boards, and all participants provided informed consent. No new individual-level data were collected for the present study; therefore, additional ethical approval was not required.

